# Public Preferences for Social Distancing Behaviors to Mitigate the Spread of COVID-19: A Discrete Choice Experiment

**DOI:** 10.1101/2020.12.12.20248103

**Authors:** Ingrid Eshun Wilson, Aaloke Mody, Ginger McKay, Mati Hlatshwayo, Cory Bradley, Vetta Thompson, Dave V Glidden, Elvin H Geng

## Abstract

Policies to promote social distancing can minimize COVID-19 transmission, but come with substantial social and economic costs. Quantifying relative preferences of the public for such practices can inform policy prioritization and optimize uptake. We used a discrete choice experiment (DCE) to quantify relative “utilities” (preferences) for five COVID-19 pandemic social distances strategies (e.g., closure of restaurants, restriction of large gatherings) against the hypothetical risk of acquiring COVID-19 and anticipated income loss. The survey was distributed in Missouri in May-June, 2020. We applied inverse probability sampling weights to mixed logit and latent class models to generate mean preferences and identify preference classes. Overall (*n*=2,428), the strongest preference was for the prohibition of large gatherings, followed by preferences to keep outdoor venues, schools, and social and lifestyle venues open, 75% of the population showing probable support for a strategy that prohibited large gatherings and closed lifestyle and social venues. Latent class analysis, however revealed four preference sub-groups in the population - “risk eliminators”, “risk balancers”, “altruistic” and “risk takers”, with men twice as likely as women to belong to the risk-taking group. In this setting, public health policies which as a first phase prohibit large gatherings, as well as close social and lifestyle venues may be acceptable and adhered to by the public. In addition, policy messages that address preference heterogeneity, for example by targeting public health messages at men, could improve adherence to social distancing measures and prevent further COVID-19 transmission prior to vaccine distribution and in the event of future pandemics.

**Significance Statement:** Preferences drive behavior – DCE’s are a novel tool in public health that allow examination of preferences for a product, service or policy, identifying how the public prioritizes personal risks and cost in relation to health behaviors. Using this method to establish preferences for COVID-19 mitigation strategies, our results suggest that, firstly, a tiered approach to non-essential business closures where large gatherings are prohibited and social and lifestyle venues are closed as a first phase, would be well aligned with population preferences and may be supported by the public, while school and outdoor venue closures may require more consideration prior to a second phase of restrictions. And secondly, that important distinct preference phenotypes - that are not captured by sociodemographic (e.g., age, sex, race) characteristics - exist, and therefore that messaging should be target at such subgroups to enhance adherence to prevention efforts.

## Introduction

In the absence of a vaccine or highly efficacious treatment, non-pharmaceutical means of stemming the COVID-19 epidemic remain necessary considerations for the foreseeable future. Many of these practices, however, carry formidable economic and social costs, giving rise to complicated considerations about potential benefits and harms for individuals as well as society at large. In the setting of at least some uncertainty, how individuals weigh the desirability and harms of such policies can help policy design to best meet public preferences when possible. Alignment with preferences will enable better uptake and greater effectiveness at stemming epidemic spread. At this moment in the COVID response, when risk of infection is rising, and calls for the intensification of social distancing policies are re-emerging, quantifying such preferences, how they group, and associated sociodemographic factors is particularly urgent.

At present, information about public views on social distancing have been prominent in the lay press. The public’s attention has been drawn toward high-profile instances where vociferous opposition to social distancing policies led to threats against public health officials. The prevalence and strength of these beliefs in the public, however, is hard to tell from media. In addition, acceptance or rejection of various distancing activities have become signatures of a particular political persuasions, but quantification is not known. Surveys published to date indicate general support for necessary measures: a study in three US urban centers found a large majority supports a range of social distancing policies (1). Surveys, however, do not capture the strength of such support and nor willingness to trade between different preferences, limiting utility for priority setting when no single solution will be sufficient, and all come with costs and harms.

In this study, we use a discrete choice experiment (DCE) – widely used in marketing – to capture preferences related to social distancing. Based in economic rational utility theory, a DCE assumes that individuals constantly seek to maximize utility (i.e., satisfaction or happiness) through deciding between different goods or services when costs prohibit having all. In a DCE respondents are offered a series of choices between two versions of a good or service where features are altered each time. Choices demonstrate the values a respondent places on a particular feature and relative to other characteristics. We have previously used DCE’s to assess preferences for HIV services (2, 3). We now apply this technique to examine preferences for social distancing measures in Missouri, a state which demographically and economically represents the US Midwest and Southern regions – areas experiencing a resurgence of new COVID cases in the fall of 2020.

## Results

Overall, 2428 respondent completed the survey between May 21, 2020 and June 13, 2020. 69% of respondents were female, 89% were White; the majority (78%) had an annual household income of greater than $50,000 and were over the age of 35 years (75%). Comorbidities were reported in 30% of respondents, 14% had an underlying chronic respiratory conditions such as asthma or other chronic lung diseases and 19% reported other comorbidities including diabetes mellitus, cancer, chronic kidney disease and other immunosuppressive disorders (Table 1). The 617 respondents who did not complete the survey were similar to completers with regard to gender and race distribution, non-completers however appeared to be older in age (S1 Table).

**Table 1:** Demographic characteristics of participants

| Demographic factor |  | Completed survey (N=2428) |
| --- | --- | --- |
| Age | 18-24yrs | 126 (6%) |
|  | 25-34yrs | 424 (19%) |
|  | 35-49yrs | 553 (25%) |
|  | 50-64yrs | 647 (29%) |
|  | 65yrs+ | 469 (21%) |
| Gender | Male | 667 (30%) |
|  | Female | 1536 (69%) |
|  | Non-conforming/other | 12 (1%) |
|  | No Answer | 4 (<1%) |
| Race | Black | 127 (6%) |
|  | White | 1973 (89%) |
|  | Other | 92 (4%) |
|  | No answer | 27 (1%) |
| Comorbidities* | No comorbidities | 1535 (69%) |
|  | Respiratory comorbidities | 320 (14%) |
|  | Other comorbidities | 431 (19%) |
|  | No answer | 11 (<1%) |
| Annual Household Income | < \$20,000 | 97 (4%) |
| | \$20,000-\$49,000 | 383 (17%) |
| | \$50,000-\$99,000 | 871 (39%) |
| | \$100,000 + | 868 (39%) |
|  | No answer | 209 (9%) |

\*Comorbidities not mutually exclusive. Other comorbidities, include: diabetes mellitus, chronic kidney disease, immunosuppressive disorders and cancer.

### Main preferences

After applying population weights, the strongest relative preferences for social distancing policy measures (Fig 1; S2 Table) were for large gatherings to be prohibited (β=-1.43; 95%CI: -1.67 to 1.18,) followed by a preference for keeping outdoor recreational venues open (β=0.50; 95%CI: 0.39 to 0.61). Maintaining social and lifestyle venues (β=0.05; 95%CI: -0.08 to 0.17) and schools open (β=0.18; 0.05 to 0.30), and shorter duration of the social distancing policy (3 versus 1 months) (β=-0.16; 95%CI: -0.31 to -0.02) showed overall weaker relative preferences. Beyond social distancing measures, participants strongest overall preference was to live in a county with a low risk of COVID infection (30% versus 5% risk) (−2.89; 95%CI: -3.23 to -2.54), followed by a preference for minimal income loss in the first six months of the policy (25% versus 5% income lost) (β=-1.49; 95% CI: -1.70 to -1.29). Moderate (of 15% compared to 5%) income loss (β= -0.72; 95% CI:-0.86 to -0.57) and risk of infection (β = -1.02; 95%CI: -1.19 to -0.84) showed relatively lower utilities.

**Figure 1:**
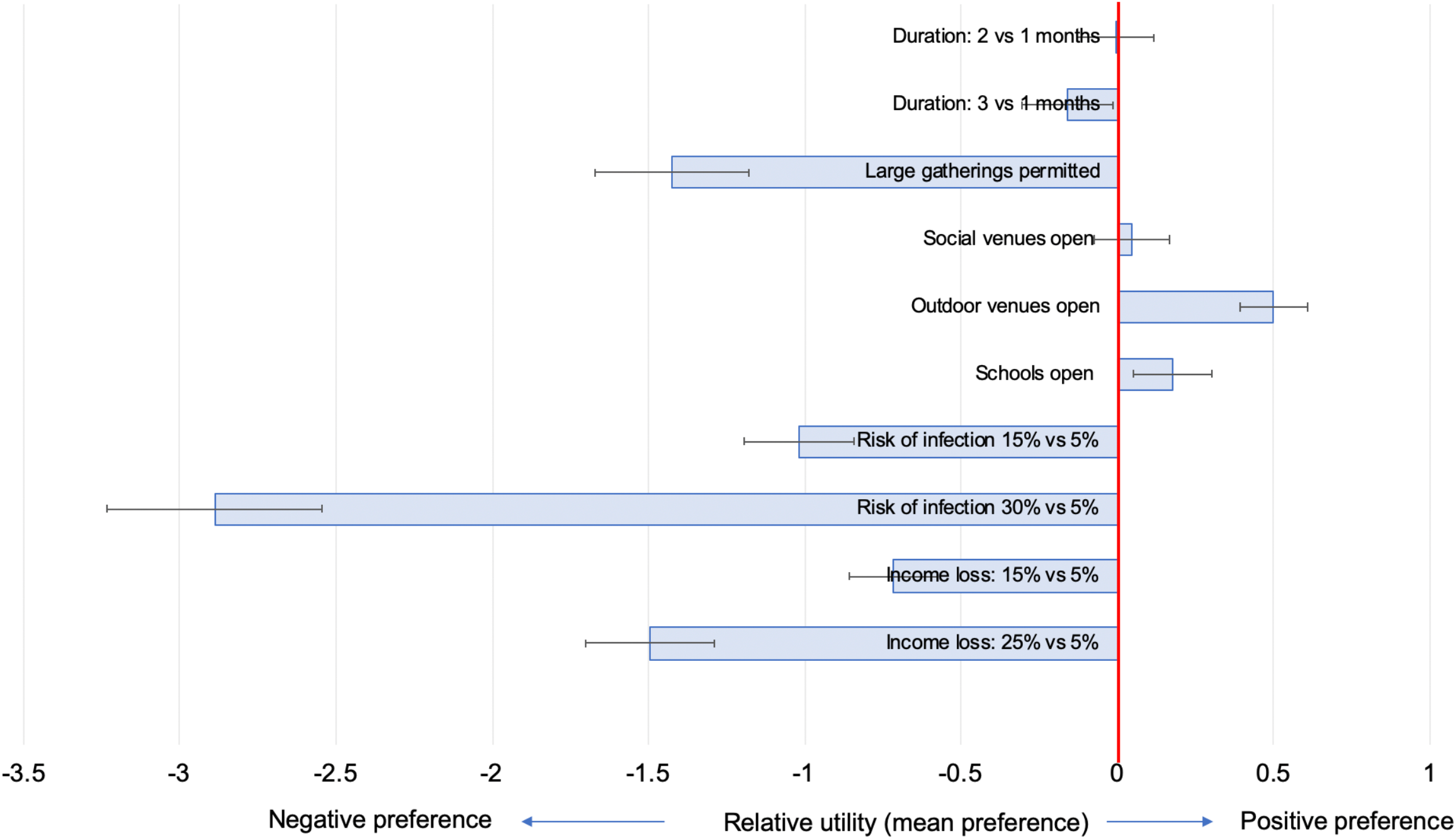
Main relative utilities (mean preferences) for social distancing policy features (N=2,428) Footnotes: main utilities and 95% confidence interval presented.

### Probability of social distancing policy uptake

Based on the main preferences for social distancing policy attributes, simulated social distancing policy scenarios show that if infection risk and income loss remain moderate (15%), a three-month social distancing policy where large gatherings such as conferences and sports event are prohibited would be preferred by 77% of the population, this preference share reduced marginally with increasing closures, with 68% of population preferring a scenario where all services (large gatherings, schools, social venues and outdoor venues) are closed. The largest drop in preference share was with increasing income loss, with the preference share dropping to 60% when income loss increased to 25% (Figure 2).

**Figure 2:**
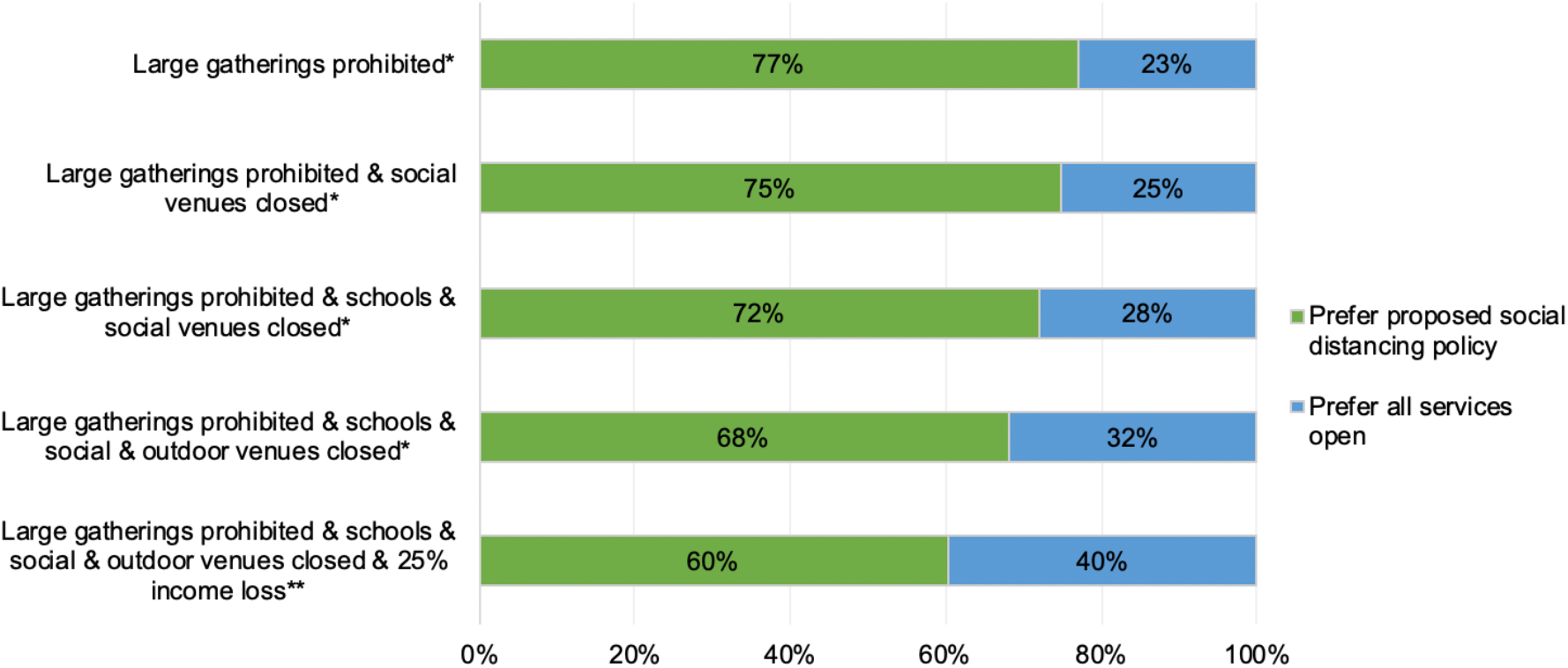
Simulated probabilities of choosing a county with varying service closures. Footnotes: Preference shares from randomized first choice model in Sawtooth Software. Preference share represents the proportion of the population who would prefer the option proposed (green) versus keeping all services open (blue). *Simulations set to a three-month policy duration, 15% income loss, 15% COVID infection risk across all groups. **Simulation set to three-month policy duration and 15% COVID infection risk for both groups, “prefer all services open” group set to 15% income loss.

**Figure 3:**
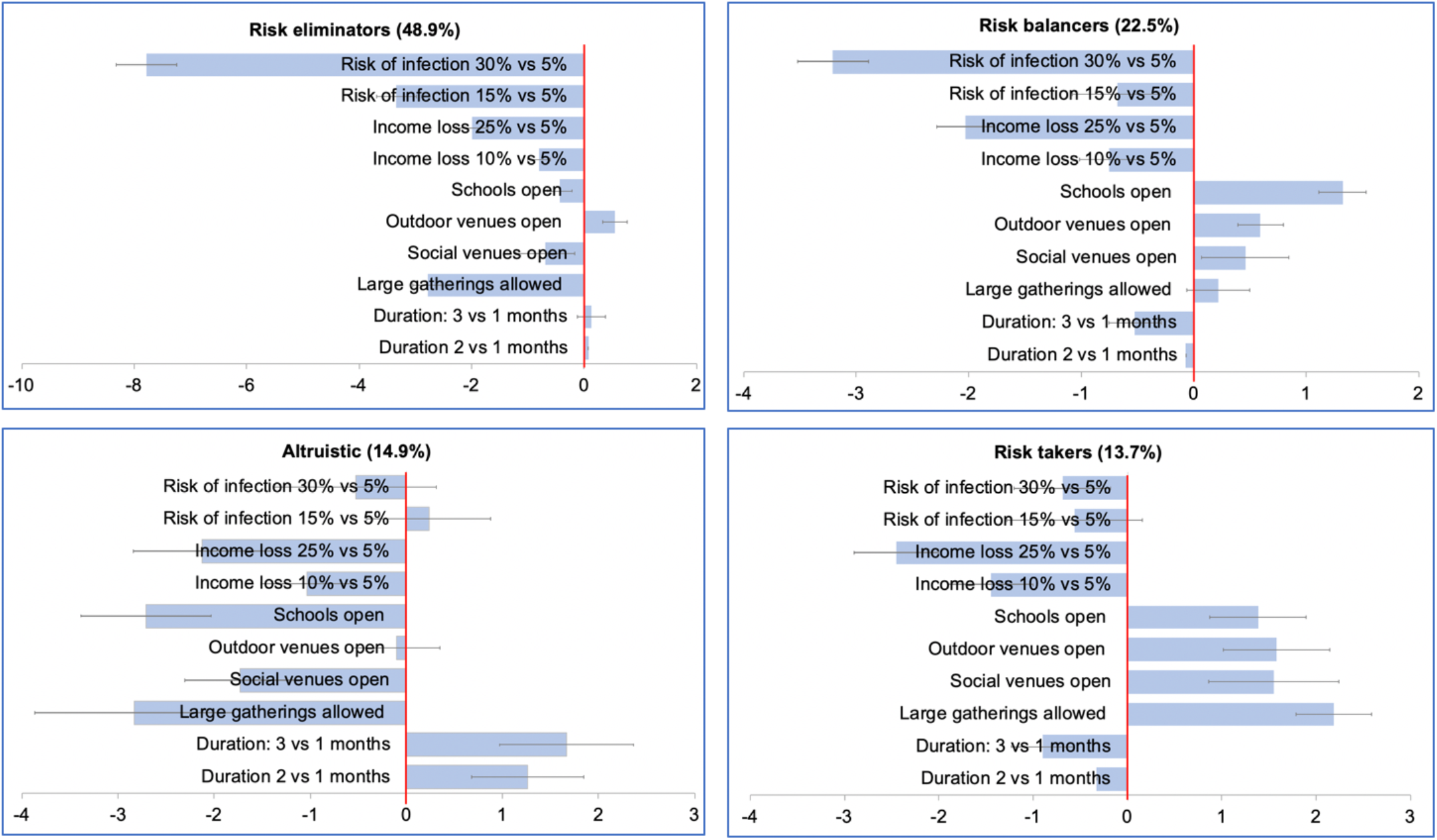
Utilities across four latent class preference groups. Four latent class groups were identified – risk eliminators, risk balancers, altruistic and risk takers. Footnotes: Utilities with 95% confidence intervals presented on x-axis, positive utilities represent positive preferences, negative utilities represent negative preferences.

### Subgroup analyses

Preferences across subgroups, largely mirrored main preferences and although some variations were seen within specific group (Table 2), there were no statistically significant differences between sub-groups across all analyses (S3-S6 Tables). Stratification by age group, however suggested that those in the youngest age category (18-24 years) showed stronger relative preferences for keeping schools and social venues open, and to prevent income loss, compared to other age groups. Men appeared to have slightly stronger preferences for maintaining outdoor recreational facilities and social/lifestyle venues open, permitting large gatherings, minimizing income loss and policy durations of 1 instead of 3 month compared to women. Stratifying by race group indicated that overall whites appeared to have stronger preferences for keeping services open (large gatherings permitted, social venues and lifestyle venues open) and shortening the duration of the policy and reducing income loss. There was no consistent difference in utilities by income category, few participants contributed to the lowest income group of <$20,000 per year annual household income.

**Table 2:** Utilities by sub-group

|  |  | Duration of<br>policy (2 vs<br>1 months) | Duration of<br>policy (3 vs<br>1 months) | Large<br>gatherings<br>permitted | Social<br>venues<br>open | Outdoor<br>venues<br>open | Schools<br>open | Risk of<br>infection<br>(15% vs 5%) | Risk of<br>infection<br>(30% vs 5%) | Percentage<br>income lost<br>(15% vs 5 %) | Percentage<br>income lost<br>(25% vs 5%) |
| --- | --- | --- | --- | --- | --- | --- | --- | --- | --- | --- | --- |
| Gender | Male | 0.01 | -0.34 | -1.31 | 0.21 | 0.61 | 0.18 | -1.06 | -3.29 | -0.88 | -1.70 |
|  | Female | 0.09 | 0.01 | -1.59 | -0.07 | 0.52 | 0.17 | -1.06 | -3.15 | -0.66 | -1.53 |
| Age | 18-24 yrs | -0.64 | -0.36 | -1.92 | -0.08 | 0.69 | 0.55 | -1.15 | -3.96 | -1.16 | -2.44 |
|  | 25-34 yrs | 0.10 | -0.31 | -1.29 | -0.03 | 0.56 | 0.10 | -1.34 | -4.12 | -0.65 | -1.79 |
|  | 35-49 yrs | 0.20 | 0.00 | -1.41 | 0.00 | 0.49 | 0.22 | -1.11 | -3.15 | -0.82 | -1.69 |
|  | 50-64 yrs | 0.04 | -0.28 | -1.22 | 0.25 | 0.66 | 0.18 | -1.14 | -3.47 | -0.85 | -1.59 |
|  | 65+ yrs | 0.06 | 0.03 | -2.24 | 0.01 | 0.56 | 0.23 | -1.31 | -3.38 | -0.67 | -1.56 |
| Race | White | -0.02 | -0.21 | -1.35 | 0.10 | 0.59 | 0.22 | -1.13 | -3.37 | -0.72 | -1.61 |
|  | Black | -0.85 | -0.40 | -5.25 | -1.24 | 0.27 | 0.21 | -1.76 | -6.87 | -1.94 | -3.39 |
|  | Other | -0.62 | 1.31 | -7.28 | -2.13 | 1.49 | -0.97 | -4.06 | -14.46 | -3.83 | -4.44 |
| Annual<br>household<br>income | < \$49,999 | 0.07 | -0.12 | -1.49 | 0.09 | 0.54 | 0.05 | -1.19 | -2.93 | -0.68 | -1.31 |
| | \$50,000-\$99,999 | 0.00 | -0.28 | -2.02 | 0.06 | 0.54 | 0.14 | -1.02 | -2.93 | -0.78 | -1.78 |
| | \$100,000 + | -0.04 | -0.21 | -1.22 | 0.10 | 0.69 | 0.39 | -1.23 | -3.68 | -0.84 | -1.81 |
| Comorbid<br>illness | Present | 0.12 | 0.12 | -2.02 | -0.24 | 0.48 | -0.05 | -1.16 | -3.44 | -0.92 | -1.77 |
|  | None | 0.02 | -0.24 | -1.33 | 0.08 | 0.65 | 0.31 | -1.19 | -3.42 | -0.69 | -1.67 |

Footnotes: Green represents negative preferences and red represents positive preferences; full data for sub-group analyses are presented in Appendix 3; there was no statistically significant difference between utilities between subgroups for all sub-group analyses.

### Latent class analysis

Four latent preference classes were identified (Figure 2, S7 Table). The largest group - “the risk eliminators” - had strong preferences for all possible restrictive policy options including the prohibition of large gatherings and keeping schools and indoor social venues closed, and a very strong preference for reducing personal COVID-19 infection risk (48.9% of the population). A second - “risk balancer” - group showed a preference for minimizing risk somewhat while balancing that risk with less restrictive policies like keeping schools open, and relatively milder preferences for keeping social and outdoor venues open, while also preferring a low personal risk of COVID infection (22.5% of the population). A third latent class group (“the altruistic”), while strongly preferring that all services remain closed and large gatherings be prohibited, showed mild preferences for reducing their personal risk of COVID-19 infection (14.9% of the population). The fourth latent class group - “the risk-takers” - showed a strong preference for keeping all services open and permitting large gatherings, with a relatively weak overall preference for reducing their COVID-19 infection risk (13.7%). Analyses of predictors of latent class membership, showed overall limited associations with individual patient preferences, except for male gender, which was a strong predictor of belonging to “the risk-takers” latent class group rather than the “risk eliminators group (S8 Table) (RRR 2.19; 95%CI: 1.54-3.12), men had a 16.7% (95%CI: 13.1-20.3%) probability of belonging to this group while women had an 8.7% (95%CI: 7.0-10.4%) probability. This trend was seen consistently across strata of income, race, comorbid illness and age (except among those over 65 years), (Figure 4).

**Figure 4:**
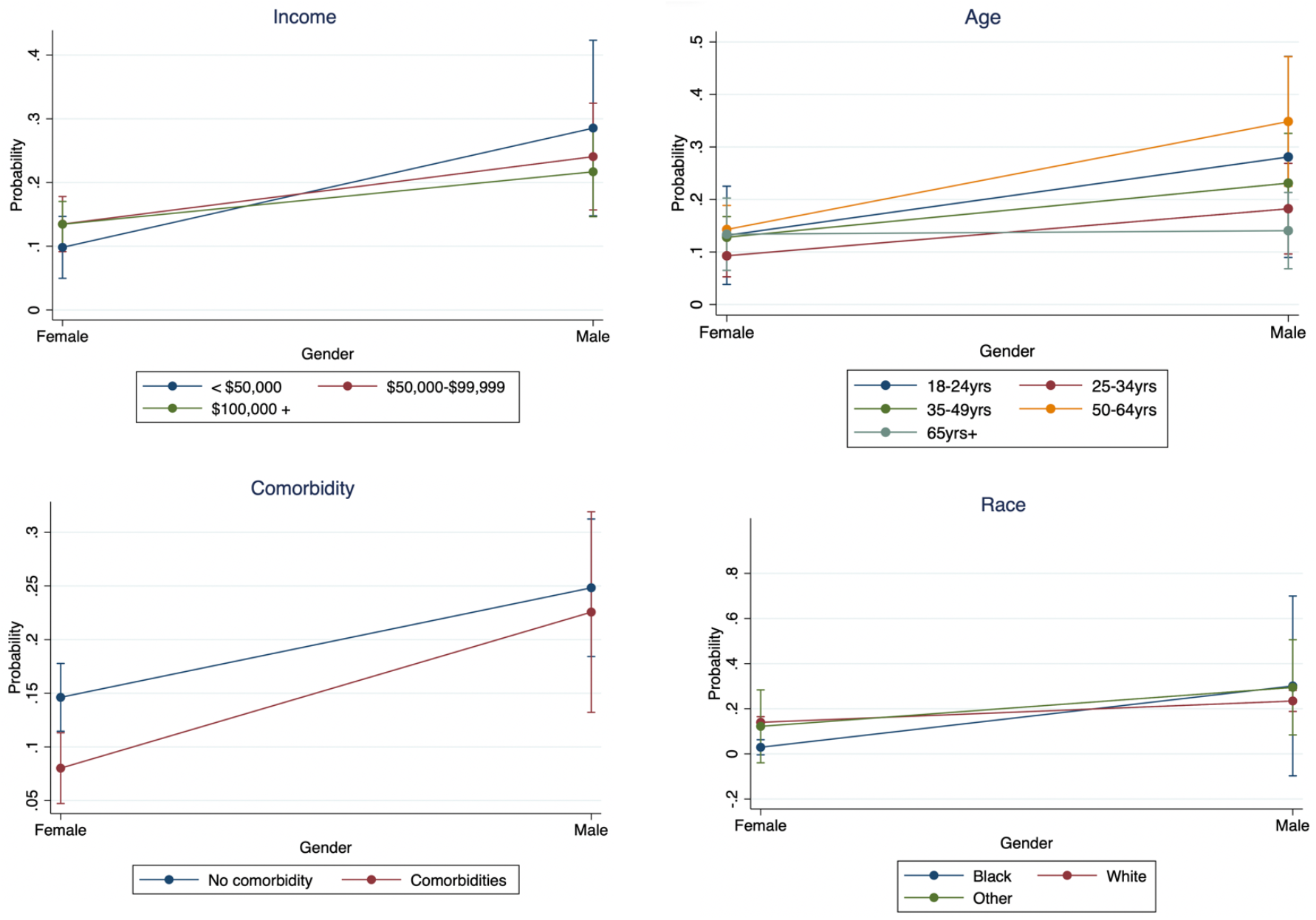
Marginal probabilities of belonging to “risk-taker” group compared to “risk eliminator” group by gender. Interactions between gender and income, age, race and comorbidity categories with 95% confidence intervals.

### Willingness to trade analysis

An analysis of trade-offs demonstrated that “risk eliminators” were willing to trade all their social freedoms (keeping schools and social / lifestyle and outdoor venues closed and prohibiting large gatherings) for a period of 3 months, in addition to giving up 25% of their income for six months (and potentially more freedoms) - to live in a county with a 5% risk of COVID infection rather than a 30% risk of infection (utility difference = -1.46; 95% CI: -2.44 to -0.47, p=0.004).

In contrast, in a willingness to risk infection analysis, the “risk taking group” were willing to tolerate a 30% risk of COVID-19 infection compared to a 5% risk (utility difference = -0.86; 95% CI: -2.07 to 0.36) to prevent losing 25% of their income for a six-month period and to maintain all services open (schools, social, lifestyle and outdoor venues and large gatherings) for a period of three months.

## Discussion

The majority of respondents in this study were markedly concerned about their risk of COVID-19 infection risk and were willing to give up a number of social freedoms and services, and a moderate percentage of their income to reduce this risk. Preferences indicated that up to 75% of the population may support a short term (3-month) social distancing policy in their community, where lifestyle and social venues including bars and salons are closed, and large gatherings such as sports events and conferences are prohibited. Latent class analysis additionally revealed four distinct preference subgroups in the community: “risk eliminators” who strongly preferred to reduce all potential COVID 19 infection risks and were willing to trade their income and all social freedoms to achieve this; the “risk balancers” who were willing to accept some COVID-19 infection risk to keep schools open; the “altruistic” who showed strong preferences for closing all services and relatively weaker preferences for reducing their own COVID-19 infection risk, and “risk-takers” who had strong preferences for keeping all services open and relatively weak preferences for reducing their own COVID risk.

Based on these data, prohibiting large gatherings such as conferences and sports events, as well as closing lifestyle and social venues including bars and salons appear to be the most acceptable service closures for the majority of the population in this setting and these preferences were evident across population subgroups. Widespread media reporting of “super-spreader events” that have fueled the COVID-19 pandemic within and outside the US likely influenced the strong preferences for the prohibition of large events in this population. In contrast the closure of social and lifestyle venues was the least important service closure relative to other policy features, indicating that, while visiting restaurants, bars, gyms and hair salons may be important, the majority of the population would be willing to trade these services to reduce their COVID-19 infection risk, and for the majority of the population maintaining schools and outdoor recreational facilities open was relatively more important than the availability of social and lifestyle services.

Latent class analysis however identified heterogenous preference subgroups, reflecting risk averse, gendered, altruistic and more balanced perspectives related to social distancing to mitigate COVID-19 transmission, and representing the heterogenous views of the pandemic and its response – heterogeneity that has implications for the segmenting of public health messaging. The largest group, comprising approximately half of the population – the risk eliminators - showed a marked aversion to COVID-19 infection risk and their preferences supported all strategies to reduce transmission – they were willing to forego or trade all social and in-person educational activities and give up a substantial proportion of their income to reduce COVID-19 infection risk in their community, supporting closure of all services for up to a three-month period. Cross-sectional surveys conducted across the US similarly report a majority of respondents supporting stay at home orders and non-essential business closures (1). A second group – the risk balancers – were willing to make some trade-offs with risk to keep some services open – particularly educational facilities. This group likely reflects the reality that while many wish to reduce their risk of COVID-19 infection, personal and financial costs of keeping services closed frequently outweigh concerns for infection risk. School closures remain a topic of debate, with marked uncertainty regarding the impact of school closures on reducing COVID-19 transmission and mortality, uncertainty reflected in population preferences in these data (4). A third preference group – the altruistic – supported the closure of all services but had relatively little concern for their own risk of COVID-19 infection. Those who have more altruistic concerns have been shown to be more likely to adhere to social distancing measures (5), urging populations to act for the common good can drive social identity, social influence and moral behavior, a potential strategy to encourage adherence to COVID-19 preventative measures in the future (6, 7). The fourth preference group – the risk-takers – (who were twice as likely to be men as compared to women) primarily focused on preserving income and keeping all social, lifestyle and educational services open, and permitting large gatherings such as conferences and sports events, with weak relative preferences for a low COVID-19 infection risk. Across strata of age, race, income and comorbid illness, men were more likely to fall into this “risk taking” group than women. A gender differential in COVID-19 risk perception and social distancing adherence, has been identified in several surveys, including cross-sectional data from over 20,000 participants across eight countries, with women more likely to comply with public health measures to prevent COVID-19 infection and to perceive COVID-19 as a very serious health problem (1, 8, 9). There were still however a proportion of women who belonged to this ‘risk taking group’, suggesting further drivers of class membership. In the US compliance with COVID-19 preventative measures has been highly politized, with those who support the Republican party less likely to adhere to such measures as compared to those who support the democratic party (10-12). This influence of bipartisanism may have additionally influenced preferences and membership in this risk-taking class.

This study has a number of limitations: Firstly, DCE’s represent hypothetical situations that may not reflect how individuals make choices in real life, findings from this study were however supported – for example with regard to gender - by evidence from cross-sectional surveys, indicating that these findings may represent revealed preferences (1, 8, 9). We did not include mask wearing as an attribute in the DCE (there was limited data regarding the benefits of mask wearing during the study design period), however survey data from other settings show that adherence to mask wearing is well aligned with adherence to other social distancing measures (10); recruitment of study participants through social media tools such as Facebook and Instagram can result in predominantly affluent female study participants whose preferences may not be representative of inference populations (13), to account for this we applied population inverse probability sampling weights, to ensure that preferences reflected the demographic structure of the Missouri. And, given that this choice experiment was conducted early during the COVID-19 pandemic, it is possible that preferences and tolerance for service closures may have changed over time.

Preferences for social distancing measures may not translate directly into adherence behavior, the preference phenotypes identified in this study however reveal that although the majority support non-essential business closures, a proportion of the Missouri population still prefer to maintain open services where small or large gatherings may promote ongoing transmission in the face of a global pandemic, reflecting the marked variation in social distancing adherence behaviors that have impeded public health efforts to stem the spread of COVID-19 transmission. As successive waves of COVID-19 transmission increase mortality rates across the US and hospitals struggle to maintain ICU capacity, public health departments again face difficult decisions regarding which social distancing measures to institute, preference data such as these can help guide decision making. For Missouri, it appears that a tiered approach which firstly prohibits large gatherings and where non-essential indoor social and lifestyle businesses are closed, prior to the closure of schools or outdoor facilities would be most acceptable. This, combined with clear, coordinated and targeted public health messaging addressing preference heterogeneity may improve adherence to social distancing measures prior to vaccine distribution and in the event of future pandemics.

## Materials and Methods

A discrete choice experiment is a survey design that solicits “utilities” from respondents.(14) Utilities have been defined as “happiness” or “preferences” and comes from economic theory. In this framing, human decision making is seen through the lens of consumer decisions in which one seeks to maximize happiness through these consumer choices and where those choices are constrained by total costs. By alternating the features (levels) of a set of service, product or policy attributes, a DCE can quantify relative utilities (preferences) for any of the features.

### Attribute selection

Selection of social distancing policy features (attributes) for inclusion in the DCE was informed by literature review and consultation with local experts in infectious diseases, social sciences and public health. We sought to identify attributes that were unconfounded, which is to say unlikely to be both representations of an underlying common but unsolicited preference, as well as present participants with a range of response categories that was wide enough to capture significance heterogeneity in preferences but within a range where a linear relationship was considered plausible for continuous attributes. We identified several candidate social distancing policy features of importance, including: (1) the duration of the policy, (2) the clarity of the messaging regarding the policy end date, (3) the closure of, childcare services, schools and colleges, indoor lifestyle services (e.g. salons, bars), outdoor recreation services (parks, beaches), religious services and mass gatherings. In addition, we determined that risk of infection or hospitalization for the individual and others, as well as income loss were other key determinants of adherence to social distancing public health measures.

The final attribute selection was supported by emerging evidence and reviews of perceptions and experiences of social distancing and quarantine, and guidelines for DCE attribute selection (15-17).

### DCE Design

In the experiment design we sought to balance pragmatism and completeness and therefore limited the number of attributes according to DCE design guidelines (five to seven attributes) and selected those attributes which we determined to be key decision drivers and of the greatest public health policy significance during the time period. To further maximize statistical and response efficiency (avoid fatigue in respondents) we limited the number of attribute levels (<=3) and the number of prohibited attribute level combinations and limited the number of DCE questions asked of each respondent to six and opted for two policy scenarios per task. We manually removed combinations considered non sensical. The final design presented consumers with two potential counties, with different sets of policies, and sought to understand which location participants preferred, all else being equal. Each policy reflected 7 attributes related to the opening or closure of social venues, education facilities and outdoor activity services, whether large gathering were permitted, the duration of the policy, the potential income lost during the first six months after the policy was instituted and the associated underlying risk of COVID infection in the county (Table 3).

**Table 3:** DCE attributes and levels

| **DCE question: In a hypothetical situation (a situation that does not necessarily exist in real life), where two counties have different social distancing policies and consequences, and you could choose to live in either, which of these two counties would you choose to live in?** |  |  |  |
| --- | --- | --- | --- |
| **Attributes** | **Social distancing policy feature options /levels** |  |  |
| **Duration of policy** | 1 month | 2 months | 3 months |
| **Percentage of income lost in six months** | 5% | 15% | 25% |
| **Educational facilities (e.g. childcare, schools, colleges)** | Open | Closed |  |
| **Outdoor activity venues (e.g. national parks, beaches)** | Open | Closed |  |
| **Large gatherings (e.g. conferences, sports, religious events)** | Permitted | Not permitted |  |
| **Social & lifestyle venues (e.g. restaurants, bars, salons, gyms)** | Open | Closed |  |
| **Your risk of COVID infection in six months** | Low risk (5%: 1 in 100 chance) | Moderate risk (10%: 1 in 10 chance) | High risk (30%: 3 in 10 chance) |

To achieve statistical efficiency, we constructed a near balanced (i.e., each level appears equally often across the experiment) and near orthogonal (i.e., each pair of levels across attributes appears equally) design using Sawtooth software and tested the design efficiency using the logit efficiency test with simulated data to obtain an efficient design with standard errors of 0.05 or less for the main-effects analysis (S9 Table). We based our sample size calculation on the formula N ≥ (500 x c)/(a x t) - where N is the number of participants, t is the number of choice tasks (questions), a is the number of alternative scenarios and c is the largest number of attribute levels for any one attribute, and when considering two-way interactions, ‘c’ is equal to the largest product of levels for any two attributes - (500 x 9/ 2 x 6) (18). To additionally conduct subgroup analyses at least 200 participants per subgroup is recommended. The DCE was powered to detect main effects, two-way interactions and evaluate at least 3 subgroups (minimum calculated sample size of 600). We followed the Professional Society for Health Economics and Outcomes Research (ISPOR) guidelines for design of choice experiments (19, 20).

### Setting and recruitment

The DCE was conducted in Missouri, a Mid-Western state in the US, with a population of 6,137,428. The majority of the population is white (83%) and 12% is Black/African American (21, 22). We used randomly allocated social media advertising on Facebook and Instagram to recruit participants in the state. In addition, the survey was distributed via email to healthy study volunteer networks, and to obtain preferences of Black/African Americans the survey was distributed through targeted social media networks linked to the Washington University Center for Community Health Partnership and Research at Washington University in St. Louis. Survey fielding commenced on 21 May 2020, a period following the lifting of a state wide stay at home order - all businesses were reopened in Missouri in early May 2020 with social distancing requirements, and full restrictions were lifted on 16 June 2020.

### Measurements

We carried out one round of cognitive interviews and piloted the final survey questions iteratively to ensure intelligibility and coherency. The survey was programmed using Sawtooth Software and participants completed the survey using personal mobile devices or computers (S10 Table). Participants were randomly allocated to one of 300 versions of the choice experiment and the order of the attributes within each question was randomized.

### Analyses

We tabulated the demographic characteristics of participants and conducted mixed logit regression to determine main utilities (preferences) for social distancing policy features. In the main effects analysis positive utilities represent a positive preference and negative utilities represent a negative preference. For all analyses we weighted responses using inverse probability weights to represent the target population of Missouri by age, gender and race (21, 22) (S11 Table). For example, if 6 % of respondents were Black/African American but this was 12% of the Missouri population, the responses of Black/African Americans were given approximately two-fold weights of whites.

To explore preference heterogeneity, we conducted sub-group analyses by gender, annual household income, age, comorbid illness and race group, and conducted latent class analysis. We fit latent class conditional logit models and used model fit criterion (Akaike and Bayesian information criterion) as well as qualitative exploration to determine the optimum number of latent classes. We validated latent class membership using cross-validation techniques (23). We used multinomial logistic regression models and relative risk ratios with 95% confidence intervals to evaluate predictors of belonging to latent classes. To compare determine marginal probabilities of belonging to one of two specific latent classes according to demographic characteristics we used generalized linear models with a log-link function.

In order to quantify trade-offs, we additionally conducted willingness to trade analyses for social distancing policies (against the percentage of income lost or risk of COVID infection in the county). Traditional willingness to pay analyses rely on the assumption of a linear relationship between continuous variables and utilities. Given that a linear relationship for infection risk and income loss could not be examined beyond the thresholds presented in the experiment, we calculated trade-offs using nonlinear combinations of estimators to determine which combination of attribute utilities were equivalent to percentage income loss or infection risk. We used Stata Version 16 to conduct mixed logit, multinomial logit and generalized linear regression analyses.

To simulate the share preference for a county/social distancing policy scenario we varied service closures using a random first choice model in Sawtooth choice simulator software. The random first choice model estimates the probability of choosing a policy or county using a simulated policy and determining the share of preference for the policy. The share preferences are presented as the percentage of the population who prefer one product compared to another. We applied this method to weighted data to determine the percentage of the Missouri population who would prefer social distancing policies with varying closure levels compared to keeping all services open.

## Data Availability

Please contact the primary author for access to the data

