## Supplementary materials for "Public Preferences for Social Distancing Behaviors to Mitigate the Spread of COVID-19: A Discrete Choice Experiment"

**This PDF file includes:**

Supplementary tables: S1 – S11

**S1 Table: Comparison of survey completers and non-completers**

| Demographic factor | | Completed survey (N=2428) | Not completed (N=617) | p-value |
|---|---|---|---|---|
| Age | 18-24yrs | 126 (6%) | 17 (4%) | <0.001 |
| | 25-34yrs | 424 (19%) | 52 (13%) | |
| | 35-49yrs | 553 (25%) | 65 (17%) | |
| | 50-64yrs | 647 (29%) | 133 (34%) | |
| | 65yrs+ | 469 (21%) | 125 (32%) | |
| Gender | Male | 667 (30%) | 130 (34%) | 0.390 |
| | Female | 1536 (69%) | 248 (65%) | |
| | Non-conforming/other | 12 (1%) | 3 (1%) | |
| | No Answer | 4 (<1%) | 1 (<1%) | |
| Race | Black | 127 (6%) | 23 (6%) | 0.610 |
| | White | 1973 (89%) | 336 (89%) | |
| | other | 92 (4%) | 12 (3%) | |
| | No answer | 27 (1%) | 7 (2%) | |
| Comorbidities | No comorbidities | 1535 (69%) | 245 (69%) | 0.84 |
| | Respiratory comorbidities | 320 (14%) | 39 (7%) | <0.001 |
| | Other comorbidities | 431 (19%) | 78 (14%) | 0.005 |
| | No answer | 11 (<1%) | 196 (36%) | <0.001 |
| Income | < $20,000 | 97 (4%) | 23 (6%) | 0.046 |
| | $20,000-$49,000 | 383 (17%) | 72 (20%) | |
| | 50,000-$99,000 | 871 (39%) | 150 (41%) | |
| | $100,000 + | 868 (39%) | 117 (32%) | |
| | No answer | 209 (9%) | 69 (16%) | |

**S2 Table: Main relative utilities (preferences) for social distancing policy features (N=2,428)**

| Attribute | Utilities | p-value | Low CI | High CI |
|---|---|---|---|---|
| Duration: 2 vs 1 months | 0.00 | 0.949 | -0.13 | 0.12 |
| Duration: 3 vs 1 months | -0.16 | 0.031 | -0.31 | -0.02 |
| Income loss: 15% vs 5% | -0.72 | 0.000 | -0.86 | -0.57 |
| Income loss: 25% vs 5% | -1.49 | 0.000 | -1.70 | -1.29 |
| Large gatherings permitted | -1.43 | 0.000 | -1.67 | -1.18 |
| Social venues open | 0.05 | 0.451 | -0.08 | 0.17 |
| Outdoor venues open | 0.50 | 0.000 | 0.39 | 0.61 |
| Schools open | 0.18 | 0.005 | 0.05 | 0.30 |
| Risk of infection 15% vs 5% | -1.02 | 0.000 | -1.19 | -0.84 |
| Risk of infection 30% vs 5% | -2.89 | 0.000 | -3.23 | -2.54 |

S3 Table & Figure: Main utilities by age category

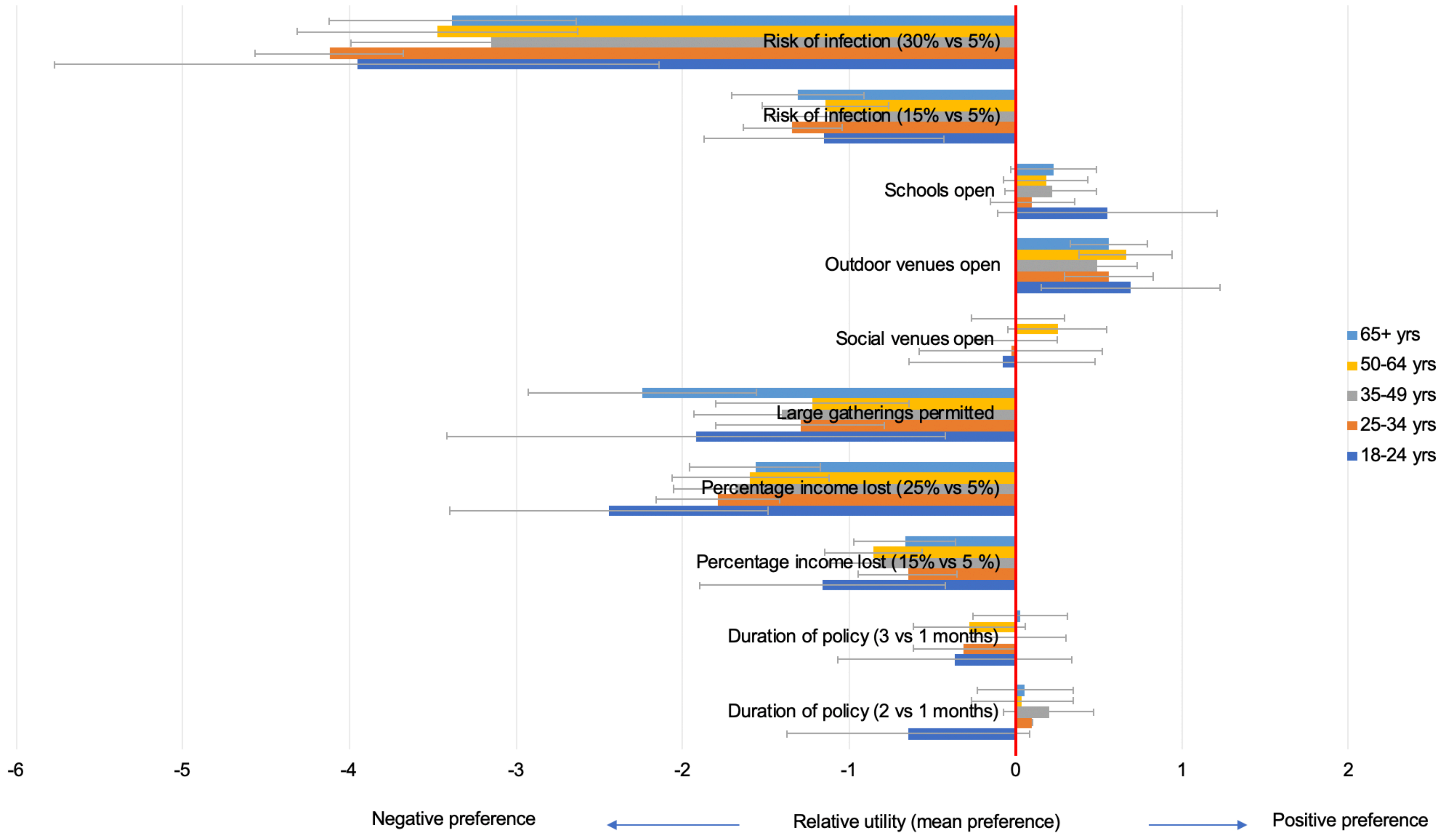

| | 18-24 yrs | | | 25-34 yrs | | | 35-49 yrs | | | 50-64 yrs | | | 65+ yrs | | |
|---|---|---|---|---|---|---|---|---|---|---|---|---|---|---|---|
| Attribute | Utility | 95% CI | | Utility | 95% CI | | Utility | 95% CI | | Utility | 95% CI | | Utility | 95% CI | |
| Duration of policy (2 vs 1 months) | -0.64 | -1.37 | 0.09 | 0.10 | -0.21 | 0.40 | 0.20 | -0.07 | 0.47 | 0.04 | -0.26 | 0.35 | 0.06 | -0.23 | 0.34 |
| Duration of policy (3 vs 1 months) | -0.36 | -1.07 | 0.34 | -0.31 | -0.61 | -0.02 | 0.00 | -0.29 | 0.30 | -0.28 | -0.62 | 0.06 | 0.03 | -0.25 | 0.31 |
| Percentage income lost (15% vs 5 %) | -1.16 | -1.90 | -0.42 | -0.65 | -1.02 | -0.27 | -0.82 | -1.12 | -0.52 | -0.85 | -1.15 | -0.56 | -0.67 | -0.97 | -0.36 |
| Percentage income lost (25% vs 5%) | -2.44 | -3.40 | -1.49 | -1.79 | -2.29 | -1.28 | -1.69 | -2.05 | -1.32 | -1.59 | -2.06 | -1.12 | -1.56 | -1.95 | -1.18 |
| Large gatherings permitted | -1.92 | -3.41 | -0.42 | -1.29 | -1.84 | -0.74 | -1.41 | -1.93 | -0.88 | -1.22 | -1.80 | -0.64 | -2.24 | -2.93 | -1.56 |
| Social venues open | -0.08 | -0.64 | 0.47 | -0.03 | -0.29 | 0.24 | 0.00 | -0.25 | 0.25 | 0.25 | -0.05 | 0.55 | 0.01 | -0.27 | 0.30 |
| Outdoor venues open | 0.69 | 0.15 | 1.23 | 0.56 | 0.31 | 0.82 | 0.49 | 0.26 | 0.73 | 0.66 | 0.38 | 0.94 | 0.56 | 0.33 | 0.79 |
| Schools open | 0.55 | -0.10 | 1.21 | 0.10 | -0.20 | 0.40 | 0.22 | -0.06 | 0.49 | 0.18 | -0.07 | 0.44 | 0.23 | -0.03 | 0.49 |
| Risk of infection (15% vs 5%) | -1.15 | -1.87 | -0.43 | -1.34 | -1.79 | -0.89 | -1.11 | -1.48 | -0.74 | -1.14 | -1.52 | -0.76 | -1.31 | -1.70 | -0.91 |
| Risk of infection (30% vs 5%) | -3.96 | -5.77 | -2.14 | -4.12 | -5.16 | -3.09 | -3.15 | -4.00 | -2.30 | -3.47 | -4.32 | -2.63 | -3.38 | -4.12 | -2.64 |

S4 Table & Figure: Main utilities by gender

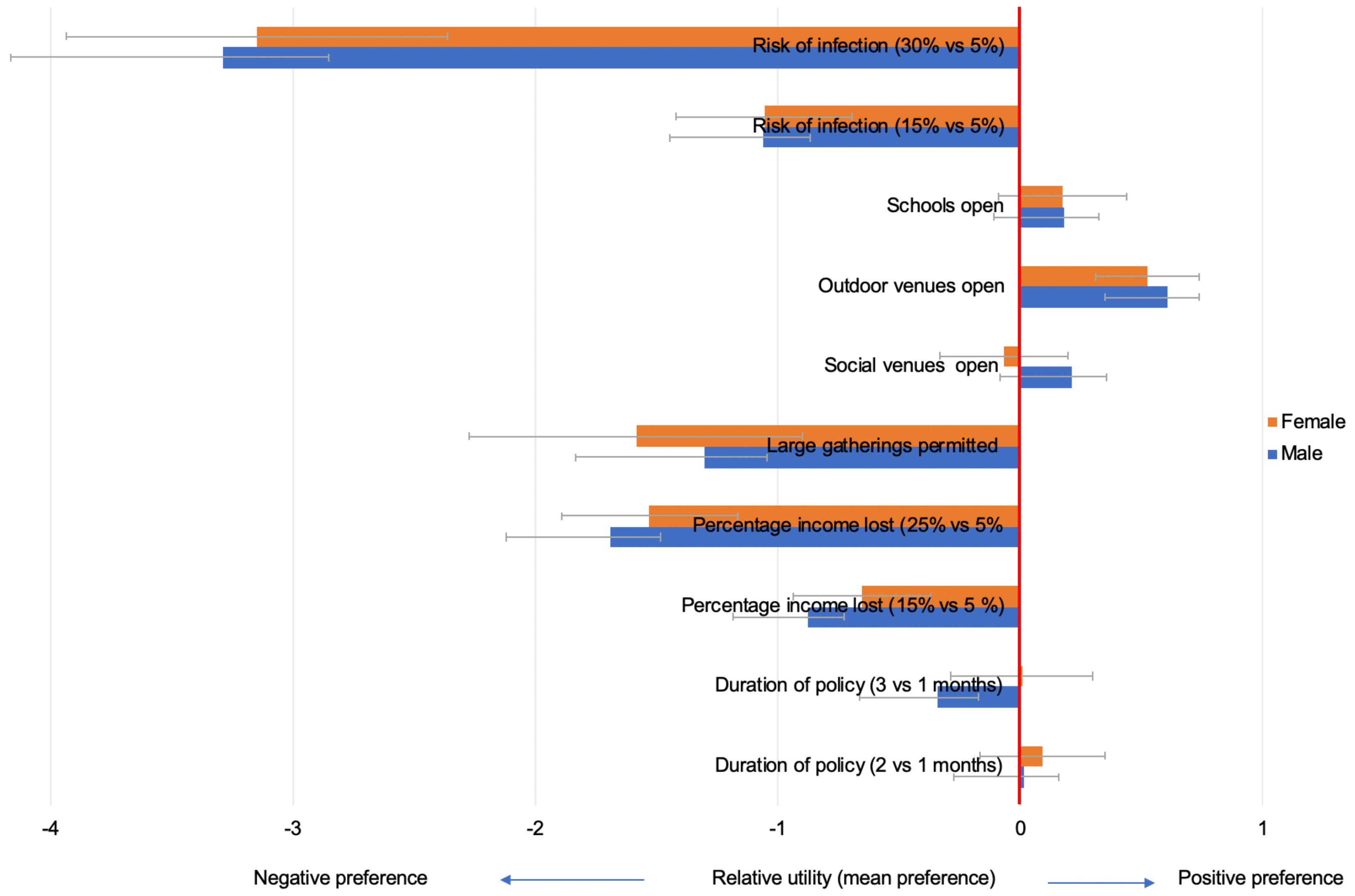

| | Male | | | Female | | |
|---|---|---|---|---|---|---|
| Attribute | Utility | 95% CI | | Utility | 95% CI | |
| Duration of policy (2 vs 1 months) | 0.01 | -0.24 | 0.27 | 0.09 | -0.05 | 0.24 |
| Duration of policy (3 vs 1 months) | -0.34 | -0.63 | -0.05 | 0.01 | -0.16 | 0.17 |
| Percentage income lost (15% vs 5 %) | -0.88 | -1.16 | -0.60 | -0.66 | -0.81 | -0.50 |
| Percentage income lost (25% vs 5% | -1.70 | -2.06 | -1.33 | -1.53 | -1.74 | -1.32 |
| Large gatherings permitted | -1.31 | -2.00 | -0.62 | -1.59 | -1.85 | -1.32 |
| Social venues open | 0.21 | -0.06 | 0.47 | -0.07 | -0.22 | 0.08 |
| Outdoor venues open | 0.61 | 0.39 | 0.82 | 0.52 | 0.39 | 0.65 |
| Schools open | 0.18 | -0.09 | 0.44 | 0.17 | 0.03 | 0.32 |
| Risk of infection (15% vs 5%) | -1.06 | -1.43 | -0.70 | -1.06 | -1.25 | -0.86 |
| Risk of infection (30% vs 5%) | -3.29 | -4.08 | -2.50 | -3.15 | -3.59 | -2.71 |

### S5 Table & Figure: Main utilities by race group

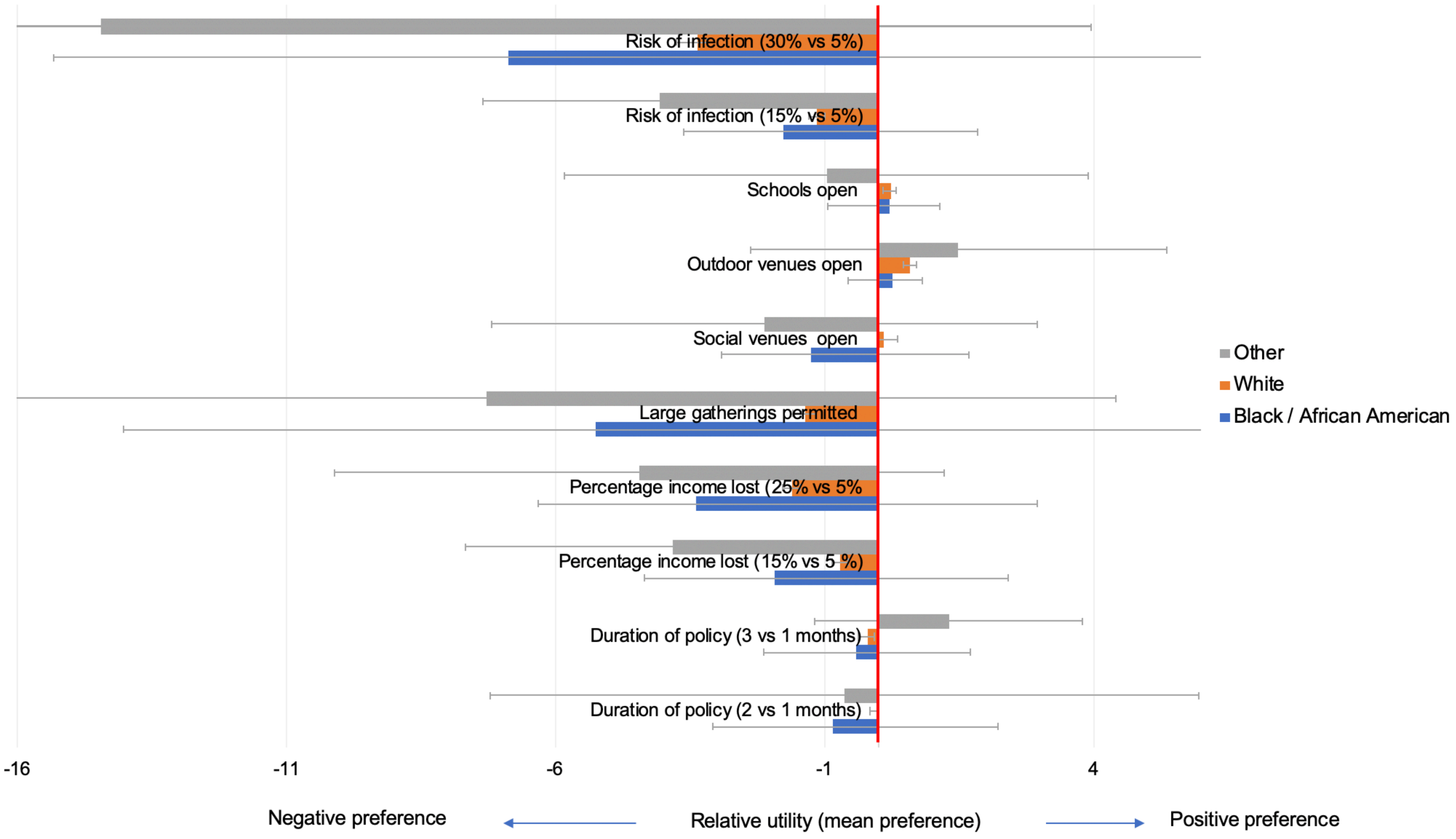

| Attribute | Black | | | White | | | Other | | |
|---|---|---|---|---|---|---|---|---|---|
| | Utility | 95% CI | | Utility | 95% CI | | Utility | 95% CI | |
| Duration of policy (2 vs 1 months) | -0.85 | -3.09 | 1.38 | -0.02 | -0.16 | 0.11 | -0.62 | -7.21 | 5.97 |
| Duration of policy (3 vs 1 months) | -0.40 | -2.13 | 1.32 | -0.21 | -0.36 | -0.06 | 1.31 | -1.17 | 3.79 |
| Percentage income lost (15% vs 5 %) | -1.94 | -4.36 | 0.49 | -0.72 | -0.86 | -0.58 | -3.83 | -7.69 | 0.02 |
| Percentage income lost (25% vs 5% | -3.39 | -6.33 | -0.45 | -1.61 | -1.79 | -1.43 | -4.44 | -10.11 | 1.24 |
| Large gatherings permitted | -5.25 | -14.05 | 3.55 | -1.35 | -1.60 | -1.10 | -7.28 | -18.98 | 4.42 |
| Social venues open | -1.24 | -2.92 | 0.44 | 0.10 | -0.02 | 0.22 | -2.13 | -7.20 | 2.95 |
| Outdoor venues open | 0.27 | -0.55 | 1.10 | 0.59 | 0.48 | 0.70 | 1.49 | -2.37 | 5.36 |
| Schools open | 0.21 | -0.94 | 1.36 | 0.22 | 0.10 | 0.35 | -0.97 | -5.85 | 3.91 |
| Risk of infection (15% vs 5%) | -1.76 | -3.62 | 0.09 | -1.13 | -1.30 | -0.96 | -4.06 | -7.36 | -0.75 |
| Risk of infection (30% vs 5%) | -6.87 | -15.34 | 1.60 | -3.37 | -3.76 | -2.98 | -14.46 | -32.88 | 3.97 |

S6 Table & Figure: Main utilities by annual household income category

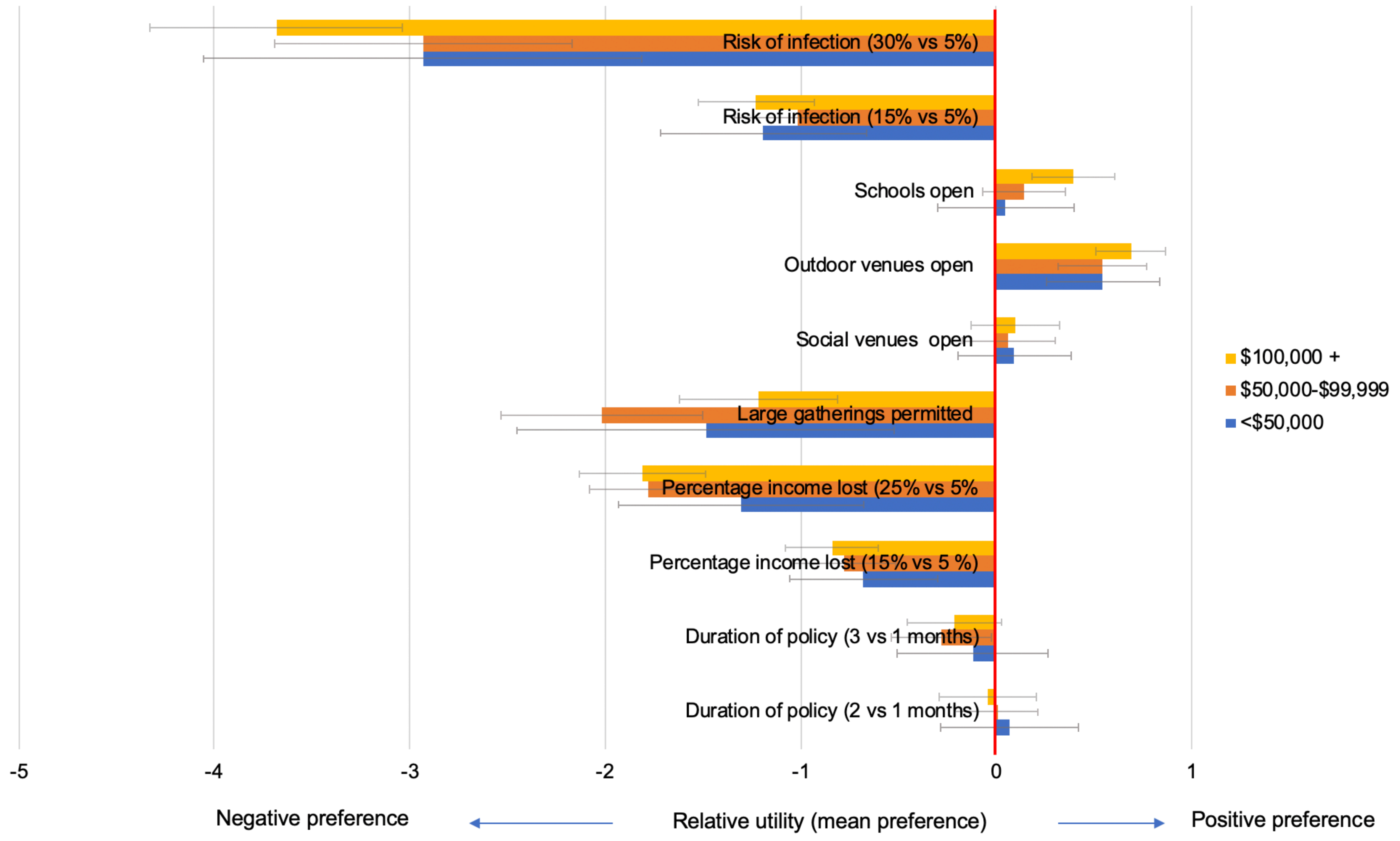

| | < $49,999 | | | $50,000-$99,999 | | | S100,000 + | | |
|---|---|---|---|---|---|---|---|---|---|
| Attribute | utility | 95%CI | | utility | 95% CI | | utility | 95% CI | |
| Duration of policy (2 vs 1 months) | 0.07 | -0.28 | 0.42 | 0.00 | -0.21 | 0.21 | -0.04 | -0.29 | 0.20 |
| Duration of policy (3 vs 1 months) | -0.12 | -0.51 | 0.26 | -0.28 | -0.54 | -0.02 | -0.21 | -0.45 | 0.03 |
| Percentage income lost (15% vs 5 %) | -0.68 | -1.06 | -0.30 | -0.78 | -1.04 | -0.52 | -0.84 | -1.08 | -0.61 |
| Percentage income lost (25% vs 5% | -1.31 | -1.94 | -0.68 | -1.78 | -2.08 | -1.48 | -1.81 | -2.13 | -1.49 |
| Large gatherings permitted | -1.49 | -2.45 | -0.52 | -2.02 | -2.53 | -1.51 | -1.22 | -1.62 | -0.82 |
| Social venues open | 0.09 | -0.19 | 0.38 | 0.06 | -0.18 | 0.30 | 0.10 | -0.13 | 0.32 |
| Outdoor venues open | 0.54 | 0.25 | 0.84 | 0.54 | 0.32 | 0.77 | 0.69 | 0.51 | 0.87 |
| Schools open | 0.05 | -0.30 | 0.40 | 0.14 | -0.07 | 0.36 | 0.39 | 0.18 | 0.60 |
| Risk of infection (15% vs 5%) | -1.19 | -1.72 | -0.67 | -1.02 | -1.34 | -0.70 | -1.23 | -1.53 | -0.93 |
| Risk of infection (30% vs 5%) | -2.93 | -4.05 | -1.81 | -2.93 | -3.69 | -2.17 | -3.68 | -4.33 | -3.04 |

S7 Table & Figure: Main utilities by comorbid illness category

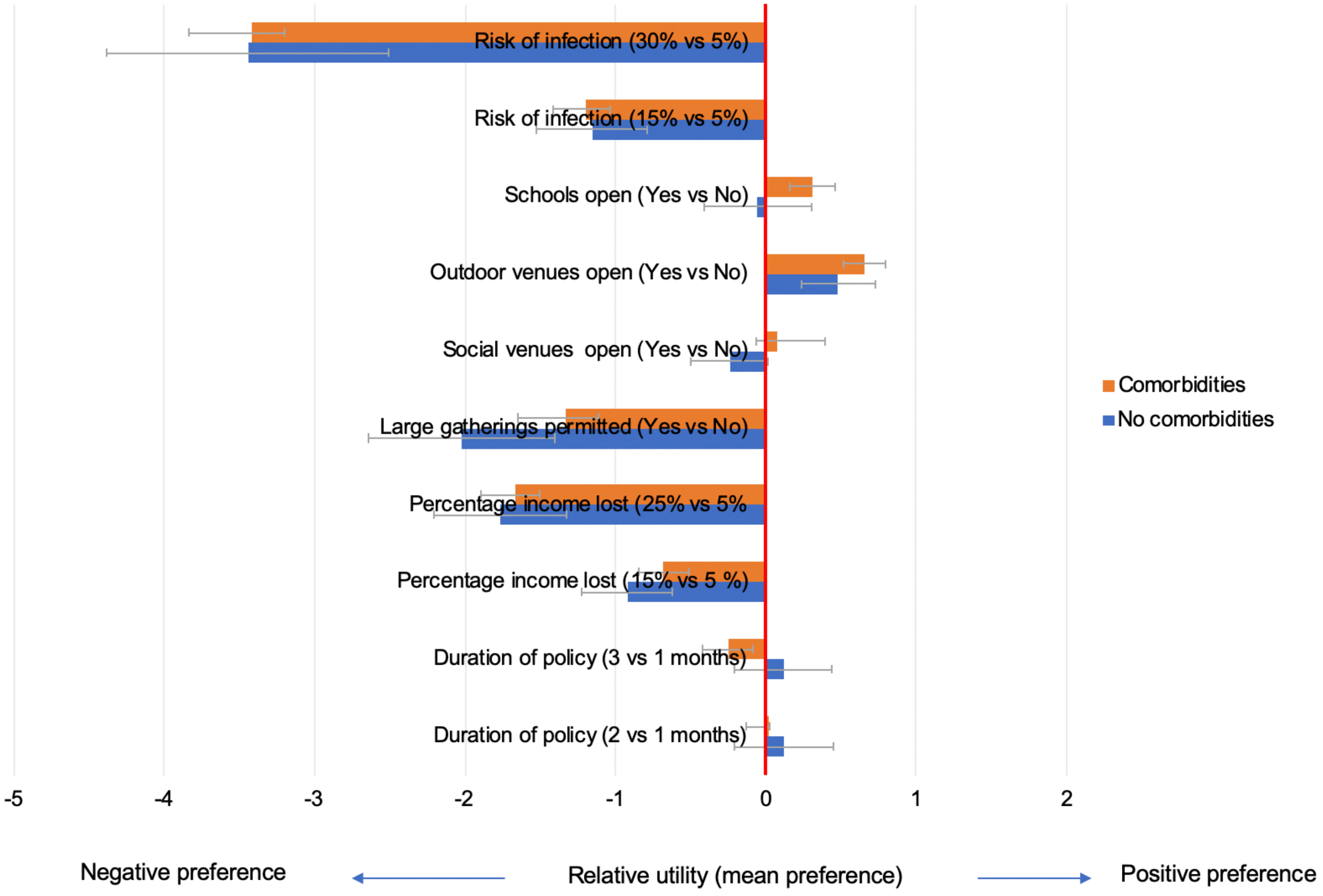

| Attribute | Comorbidity | | | No comorbidity | | |
|---|---|---|---|---|---|---|
| | Utility | 95%CI | | Utility | 95%CI | |
| Duration of policy (2 vs 1 months) | 0.12 | -0.21 | 0.44 | 0.02 | -0.13 | 0.17 |
| Duration of policy (3 vs 1 months) | 0.12 | -0.21 | 0.44 | -0.24 | -0.42 | -0.07 |
| Percentage income lost (15% vs 5 %) | -0.92 | -1.23 | -0.62 | -0.69 | -0.85 | -0.52 |
| Percentage income lost (25% vs 5%) | -1.77 | -2.21 | -1.32 | -1.67 | -1.89 | -1.45 |
| Large gatherings permitted | -2.02 | -2.64 | -1.40 | -1.33 | -1.65 | -1.02 |
| Social venues open | -0.24 | -0.49 | 0.02 | 0.08 | -0.06 | 0.22 |
| Outdoor venues open | 0.48 | 0.23 | 0.72 | 0.65 | 0.51 | 0.79 |
| Schools open | -0.05 | -0.41 | 0.30 | 0.31 | 0.16 | 0.47 |
| Risk of infection (15% vs 5%) | -1.16 | -1.52 | -0.79 | -1.19 | -1.41 | -0.98 |
| Risk of infection (30% vs 5%) | -3.44 | -4.38 | -2.51 | -3.42 | -3.84 | -3.00 |

**S8 Table: Utilities by latent class membership**

| Levels | Altruistic | | | Risk takers | | | Risk eliminators | | | Risk averse educators | | |
|---|---|---|---|---|---|---|---|---|---|---|---|---|
| | Utility | 95% CI | | Utility | 95% CI | | Utility | 95% CI | | Utility | 95% CI | |
| Duration: 2 vs 1 months | 1.26 | 0.68 | 1.84 | -0.33 | -0.66 | 0.01 | 0.08 | -0.17 | 0.33 | -0.07 | -0.30 | 0.16 |
| Duration: 3 vs 1 months | 1.67 | 0.98 | 2.36 | -0.90 | -1.30 | -0.50 | 0.13 | -0.12 | 0.39 | -0.52 | -0.80 | -0.24 |
| Income loss: 10% vs 5% | -1.04 | -1.51 | -0.56 | -1.45 | -1.90 | -1.00 | -0.80 | -1.08 | -0.52 | -0.75 | -1.01 | -0.49 |
| Income loss: 25% vs 5% | -2.12 | -2.84 | -1.40 | -2.45 | -3.17 | -1.73 | -1.99 | -2.34 | -1.65 | -2.03 | -2.43 | -1.63 |
| Large gatherings: permitted vs not | -2.83 | -3.87 | -1.80 | 2.19 | 1.50 | 2.87 | -2.78 | -3.30 | -2.27 | 0.22 | -0.17 | 0.61 |
| Social venues open vs closed | -1.73 | -2.31 | -1.16 | 1.55 | 0.99 | 2.11 | -0.69 | -0.90 | -0.47 | 0.46 | 0.26 | 0.66 |
| Outdoor venues open vs closed | -0.10 | -0.56 | 0.35 | 1.58 | 1.07 | 2.10 | 0.55 | 0.34 | 0.77 | 0.60 | 0.39 | 0.80 |
| Schools open vs closed | -2.71 | -3.39 | -2.04 | 1.38 | 0.93 | 1.84 | -0.43 | -0.63 | -0.23 | 1.33 | 1.06 | 1.59 |
| Risk of infection 15% vs 5% | 0.23 | -0.41 | 0.87 | -0.56 | -1.07 | -0.05 | -3.33 | -3.87 | -2.80 | -0.68 | -0.99 | -0.36 |
| Risk of infection 30% vs 5% | -0.53 | -1.37 | 0.32 | -0.69 | -1.46 | 0.09 | -7.77 | -8.93 | -6.62 | -3.20 | -3.84 | -2.57 |

**S9 Table: Predictors of latent class membership (multinomial logistic regression, baseline comparison group "risk eliminators")**

| Characteristic | | Risk averse educators (22.5%) | | | | Altruistic (14.9%) | | | | Risk-takers (13.7%) | | | |
|---|---|---|---|---|---|---|---|---|---|---|---|---|---|
| | | RRR | 95% CI | | p-value | RRR | 95% CI | | p-value | RRR | 95% CI | | p-value |
| Gender | Female | 1.00 | | | 0.455 | 1.00 | | | 0.660 | 1.00 | | | <0.001 |
| | Male | 1.12 | 0.84 | 1.49 | | 1.09 | 0.75 | 1.58 | | 2.19 | 1.54 | 3.12 | |
| Age | 18-24yrs | 1.00 | | | 0.317 | 1.00 | | | 0.674 | 1.00 | | | 0.057 |
| | 25-34yrs | 0.98 | 0.54 | 1.78 | | 1.37 | 0.63 | 2.94 | | 0.60 | 0.28 | 1.27 | |
| | 35-49yrs | 0.68 | 0.37 | 1.25 | | 1.60 | 0.75 | 3.40 | | 0.84 | 0.40 | 1.77 | |
| | 50-64yrs | 0.87 | 0.48 | 1.56 | | 1.58 | 0.69 | 3.58 | | 1.23 | 0.57 | 2.65 | |
| | 65yrs + | 0.71 | 0.38 | 1.33 | | 1.23 | 0.57 | 2.65 | | 0.65 | 0.29 | 1.43 | |
| Income | < $50,000 | 1.00 | | | 0.102 | 1.00 | | | 0.123 | 1.00 | | | 0.887 |
| | 50,000 < 99,000 | 1.41 | 0.93 | 2.14 | | 0.69 | 0.43 | 1.11 | | 0.96 | 0.58 | 1.61 | |
| | $100,000 + | 1.49 | 0.99 | 2.25 | | 0.70 | 0.44 | 1.10 | | 0.93 | 0.56 | 1.55 | |
| Race | White | 1.00 | | | 0.679 | 1.00 | | | 0.333 | 1.00 | | | 0.628 |
| | Black | 0.88 | 0.44 | 1.76 | | 0.77 | 0.34 | 1.72 | | 0.58 | 0.18 | 1.82 | |
| | Other | 0.76 | 0.38 | 1.49 | | 1.57 | 0.80 | 3.10 | | 1.09 | 0.48 | 2.48 | |
| Comorbidity | No | 1.00 | | | 0.066 | 1.00 | | | 0.478 | 1.00 | | | 0.194 |
| | Yes | 0.74 | 0.53 | 1.02 | | 1.14 | 0.79 | 1.64 | | 0.74 | 0.47 | 1.16 | |

Footnotes: Baseline comparison class: risk eliminators (48.9% of population)

CBC Design: Preliminary Counting Test
Copyright Sawtooth Software
10/20/2020 8:20:53 PM

Using an imported design.
Based on 300 version(s).
Includes 1800 total choice tasks (6 per version).
Each choice task includes 2 concepts and 7 attributes.

| Att | Lev | Freq | Level |
|---|---|---|---|
| 1 | 1 | 1200 | time_1 |
| 1 | 2 | 1200 | time_2 |
| 1 | 3 | 1200 | time_3 |
| 2 | 1 | 1200 | five |
| 2 | 2 | 1199 | fifteen |
| 2 | 3 | 1201 | twenty five |
| 3 | 1 | 1799 | no_gather |
| 3 | 2 | 1801 | yes_gather |
| 4 | 1 | 1800 | socialvenue_yes |
| 4 | 2 | 1800 | socialvenue_no |
| 5 | 1 | 1800 | outdoor_no |
| 5 | 2 | 1800 | outdoor_yes |
| 6 | 1 | 1800 | open |
| 6 | 2 | 1800 | closed |
| 7 | 1 | 1178 | five |
| 7 | 2 | 1245 | fifteen |
| 7 | 3 | 1177 | thirty |

**Warning: You have specified prohibitions/alternative-specific rules between two or more attributes. You cannot automatically estimate an interaction effect between two attributes when a prohibition or alternative-specific rule is in place.

**Warning: We strongly encourage you to further investigate the efficiency of your design prior to fielding this study.

Two-Way Frequencies
-------------------------------------------------------------
Att/Lev 1/1 1/2 1/3 2/1 2/2 2/3
3/1 3/2 4/1 4/2 5/1 5/2 6/1

```
6/2        7/1        7/2        7/3
1/1             1200          0          0        400        401        399
600        600        600        600        599        601        601
599        391        415        394
1/2                0       1200          0        401        398        401
600        600        599        601        600        600        601
599        394        414        392
1/3                0          0       1200        399        400        401
599        601        601        599        601        599        598
602        393        416        391
2/1              400        401        399       1200          0          0
598        602        599        601        600        600        602
598        391        418        391
2/2              401        398        400          0       1199          0
600        599        600        599        600        599        598
601        391        414        394
2/3              399        401        401          0          0       1201
601        600        601        600        600        601        600
601        396        413        392
3/1              600        600        599        598        600        601
1799          0        899        900        898        901        899
900       1178        621          0
3/2              600        600        601        602        599        600
0      1801        901        900        902        899        901
900          0        624       1177
4/1              600        599        601        599        600        601
899        901       1800          0        900        900        897
903        588        622        590
4/2              600        601        599        601        599        600
900        900          0       1800        900        900        903
897        590        623        587
5/1              599        600        601        600        600        600
898        902        900        900       1800          0        901
899        589        622        589
5/2              601        600        599        600        599        601
901        899        900        900          0       1800        899
901        589        623        588
6/1              601        601        598        602        598        600
899        901        897        903        901        899       1800
0        589        624        587
6/2              599        599        602        598        601        601
900        900        903        897        899        901          0
1800        589        621        590
7/1              391        394        393        391        391        396
1178          0        588        590        589        589        589
589       1178          0          0
7/2              415        414        416        418        414        413
621        624        622        623        622        623        624
621          0       1245          0
7/3              394        392        391        391        394        392
```

0      1177       590       587       589       588       587
590         0         0      1177

This counting test reports how balanced the design is in terms of frequencies. To assess how precisely this design can estimate utilities given your expected sample size, we recommend you refer to the Test Design report (the Logit Efficiency Test Using Simulated Data).

Logit Efficiency Test Using Simulated Data
-------------------------------------------------------------
Main Effects: 1 2 3 4 5 6 7
Build includes 600 respondents.

Total number of choices in each response category:

| Category | Number | Percent |
|---|---|---|
| 1 | 1817 | 50.47% |
| 2 | 1783 | 49.53% |

There are 3600 expanded tasks in total, or an average of 6.0 tasks per respondent.

Iter    1  Log-likelihood = -2490.30855  Chi Sq = 10.04259  RLH = 0.50070
Iter    2  Log-likelihood = -2490.10417  Chi Sq = 10.45136  RLH = 0.50073
Iter    3  Log-likelihood = -2490.09591  Chi Sq = 10.46788  RLH = 0.50073
Iter    4  Log-likelihood = -2490.09558  Chi Sq = 10.46854  RLH = 0.50073
Iter    5  Log-likelihood = -2490.09557  Chi Sq = 10.46857  RLH = 0.50073
Iter    6  Log-likelihood = -2490.09557  Chi Sq = 10.46857  RLH = 0.50073
*Converged

| | Std Err | Attribute Level |
|---|---|---|
| 1 | 0.03036 | 1 1 time_1 |
| 2 | 0.03020 | 1 2 time_2 |
| 3 | 0.03011 | 1 3 time_3 |
| 4 | 0.03028 | 2 1 five |
| 5 | 0.03042 | 2 2 fifteen |
| 6 | 0.03001 | 2 3 twenty five |
| 7 | 0.03951 | 3 1 no_gather |

```
  8       0.03951    3 2 yes_gather

  9       0.01950    4 1 socialvenue_yes
 10       0.01950    4 2 socialvenue_no

 11       0.01965    5 1 outdoor_no
 12       0.01965    5 2 outdoor_yes

 13       0.02013    6 1 open
 14       0.02013    6 2 closed

 15       0.05315    7 1 five
 16       0.02992    7 2 fifteen
 17       0.05339    7 3 thirty
```

A general guideline is to achieve standard errors of 0.05 or smaller for main effect utilities and 0.10 or smaller for interaction effects or alternative-specific effects.

The strength of design for this model is 1479.71421
(The ratio of strengths of design for two designs reflects the D-Efficiency of one design relative to the other.)

**S11 Table: Weighting strategy based on Missouri population in 2019**

| Weighting strata (race, gender, age category) | Missouri population proportion in strata | Number of DCE participants in strata | Proportion of DCE population in strata | Inverse probability weight applied to strata |
|---|---|---|---|---|
| black_fem_18_24 | 0.007 | 5 | 0.002 | 3.730 |
| black_fem_25_34 | 0.010 | 14 | 0.006 | 1.580 |
| black_fem_35_49 | 0.014 | 37 | 0.017 | 0.843 |
| black_fem_50_64 | 0.015 | 34 | 0.013 | 1.194 |
| black_fem_65 | 0.015 | 17 | 0.007 | 1.994 |
| black_male_18_24 | 0.007 | 3 | 0.001 | 7.717 |
| black_male_25_34 | 0.010 | 8 | 0.004 | 2.797 |
| black_male_35_49 | 0.014 | 10 | 0.005 | 2.984 |
| black_male_50_64 | 0.014 | 5 | 0.002 | 6.174 |
| black_male_65 | 0.012 | 2 | 0.001 | 12.605 |
| other_fem_18_24 | 0.005 | 12 | 0.005 | 1.046 |
| other_fem_25_34 | 0.008 | 17 | 0.008 | 1.004 |
| other_fem_35_49 | 0.011 | 18 | 0.008 | 1.377 |
| other_fem_50_64 | 0.012 | 11 | 0.004 | 3.224 |
| other_fem_65 | 0.011 | 6 | 0.002 | 6.150 |
| other_male_18_24 | 0.005 | 8 | 0.003 | 1.700 |
| other_male_25_34 | 0.008 | 9 | 0.004 | 1.918 |
| other_male_35_49 | 0.011 | 9 | 0.004 | 2.557 |
| other_male_50_64 | 0.011 | 6 | 0.003 | 3.968 |
| other_male_65 | 0.009 | 5 | 0.001 | 6.480 |
| white_fem_18_24 | 0.046 | 64 | 0.027 | 1.724 |
| white_fem_25_34 | 0.068 | 267 | 0.120 | 0.566 |
| white_fem_35_49 | 0.093 | 376 | 0.167 | 0.561 |
| white_fem_50_64 | 0.103 | 462 | 0.191 | 0.539 |
| white_fem_65 | 0.098 | 325 | 0.123 | 0.795 |
| white_male_18_24 | 0.047 | 40 | 0.017 | 2.723 |
| white_male_25_34 | 0.069 | 111 | 0.049 | 1.402 |
| white_male_35_49 | 0.092 | 119 | 0.050 | 1.818 |
| white_male_50_64 | 0.095 | 181 | 0.078 | 1.210 |
| white_male_65 | 0.078 | 191 | 0.078 | 0.994 |
